# Associations of Cumulative Perceived Stress with Cardiovascular Risk Factors and Outcomes: Findings from The Dallas Heart Study

**DOI:** 10.1101/2023.06.15.23291460

**Authors:** Ijeoma Eleazu, Colby Ayers, Ann Marie Navar, Karim Salhadar, Michele Albert, Mercedes Carnethon, Sherwood Brown, Lucy Ogbu Nwobodo, Spencer Carter, Courtney Bess, Tiffany M. Powell-Wiley, James A. de Lemos

**Author notes:** Corresponding Author: Ijeoma Eleazu MD, University of Texas Southwestern Medical Center 5323 Harry Hines Blvd., Dallas Texas, 75390-8830.

## Abstract

**Background:** Data remain sparse regarding the impact of chronic stress on cardiovascular disease (CVD) risk factors and outcomes. Prior work has been limited by incomplete assessments of perceived stress and focus on single stress domains. We evaluated the association between a composite measure of perceived stress and CVD risk factors and outcomes.

**Methods:** Participants from the Dallas Heart Study phase 2 (2007-2009) without prevalent CVD who completed questionnaire assessments of perceived stress were included (n=2685). Individual perceived stress subcomponents (generalized stress, psychosocial, financial, and neighborhood stress) were standardized and integrated into a single cumulative stress score (CSS) with equal weighting for each component. Associations between CSS and demographics, psychosocial variables and cardiac risk factors were assessed in univariable and multivariable analyses. Cox proportional hazards models were used to determine associations of the CSS with atherosclerotic CVD (ASCVD) and Global CVD (ASCVD, heart failure, and atrial fibrillation) after adjustment for demographics and traditional risk factors.

**Results:** Median age of the study population was 48 years, 55% were female, 49% Black and 15% Hispanic/Latinx. CSS was higher among participants who were younger, female, Black or Hispanic, and those with lower income and educational attainment (p<.0001 for each). Higher CSS was associated with self-report of racial/ethnic discrimination, lack of health insurance and last medical contact > one year previously (p<.0001 for each). In multivariable regression models adjusting for age, gender, race/ethnicity, income and education, higher CSS associated with hypertension, smoking, and higher body mass index, waist circumference Hemoglobin A1C, hs-CRP and sedentary time (p< 0.01 for each). Over a median follow-up of 12.4 years, higher CSS associated with ASCVD (adjusted HR 1.22 per SD, 95% CI 1.01-1.47) and Global CVD (HR 1.20, 95% CI 1.03-1.40). No interactions were seen between CSS, demographic factors, and outcomes.

**Conclusion:** Composite multidimensional assessments of perceived stress may help to identify individuals at risk for CVD who may be targeted for stress mitigation or enhanced prevention strategies. These approaches may be best focused on vulnerable populations, given the higher burden of stress in women, Black and Hispanic individuals, and those with lower income and education.

**WHAT IS NEW?:**

- A novel measure of cumulative stress was created that integrates generalized, psychosocial, financial, and neighborhood perceived stress.
- Cumulative stress was higher among women, Black and Hispanic participants, younger individuals and persons with lower income and educational attainment and was associated with adverse health behaviors and increased burden of cardiovascular disease (CVD) risk factors.
- In a diverse cohort, higher cumulative stress associated with incident CVD after adjustment for demographics and traditional risk factors. No interactions were seen based on demographic factors.

**CLINICAL IMPLICATIONS:**

- Although associations of chronic stress with CVD were similar across demographic subgroups, the higher burden of stress among younger individuals, women, Black and Hispanic participants, and those with lower SES suggests that CVD risk associated with higher stress affects marginalized groups disproportionately.
- Cumulative Stress is associated with modifiable risk factors and health behaviors. Future studies should explore targeting behavioral modification and risk factor reduction programs, as well as stress reduction strategies, to individuals with high cumulative stress.
- Additional research is needed to uncover mechanisms that underly the association between chronic stress and cardiovascular disease.

## Introduction

Increasing evidence suggests a link between chronic stress and cardiovascular disease (CVD) risk factors and outcomes. These associations remain despite adjustment for a higher burden of traditional CVD risk factors among those with higher perceived stress[1–3]. However, significant knowledge gaps remain regarding the impact of chronic perceived stress on development of CVD. Prior studies have largely assessed single domains of stressful experiences such as “work stress” or “marital stress”[4–6] or perceived stressful feelings rather than considering a cumulative model of stress.

Factors encompassing stress are multi-level, comprising individual, demographic, and environmental components. An individual’s psychosocial stress exposure can result from numerous insults and lead to depressive symptoms that have been associated with increased risk for CVD [7, 8]. Financial hardship is a common chronic stressor that influences access to resources that may impact health status and impair coping strategies and prognosis [9]. Residential neighborhoods are important determinants of health irrespective of neighborhood level socioeconomic status [10]. Perceptions of adverse neighborhood characteristics have been linked with CVD risk factors such as obesity and have influence on adverse cardiovascular health behaviors [11]. Therefore, the recognition of stress as a potentially modifiable risk factor mandates a conceptual model that considers a more comprehensive assessment of different components of stress[12].

Given the paucity of data about the role of cumulative stress on CVD risk, we developed a composite stress measure that integrated generalized, psychosocial, financial, and neighborhood perceived stress and determined associations with CVD outcomes in The Dallas Heart Study (DHS). The DHS is a multi-ethnic cohort with extensive available information about socioeconomic, clinical, and psychosocial factors along with detailed cardiometabolic phenotyping, thereby offering the unique opportunity to evaluate relationships between perceived stress and heart disease.

## Methods

### Study Population

The DHS is a multiethnic, population-based cohort study of residents in Dallas County. Self-Identified Black individuals were intentionally oversampled to encompass 50% of the cohort [14]. The original DHS participants were enrolled from 2000-2002. Participants and their spouses or significant others were invited to attend DHS phase 2 (DHS-2), a follow up examination performed between 2007-2009. DHS-2 included 3403 participants who underwent a comprehensive health examination including collection of extensive survey information, measurement of blood pressure, collection of blood and urine samples, cardiac and brain magnetic resonance imaging (MRI), accelerometry, and cardiac fitness testing [15].

Details of the DHS design and cohort have been reported previously [16]. For the present study, we included participants from DHS-2 who completed the questionnaire assessments outlined below and excluded participants with prevalent CVD or missing stress assessment data. The final study population included 2975 participants (see supplementary figure 1). All participants provided written informed consent and the study was approved by the University of Texas Southwestern Institutional Review Board

## Measurement of Stress

### Generalized Perceived Stress (PSS-4)

Generalized perceived stress was assessed using the 4-item version of the Perceived Stress Score (PSS). The PSS is a reliable measure of an individual’s sense of control and confidence in handling circumstances over the past month and has been validated in low literacy populations[17, 18]. The four questions ask about the degree to which respondents feel in control of important aspects in their life, their confidence in ability to handle personal problems, how often they felt things were going their way and how often difficulties were overwhelming. Participants responded to each question on a five-point scale ranging from never (1) to very often (5). Of the four items, two of the questions were positively worded and thus were reverse coded. There is no established cut off for high stress on the PSS. Responses were summed to create a total score which ranged from 4-20.

### Perceived Psychosocial Stress

The psychosocial variables: presence of depressive symptoms, lack of calm, and lack of energy were assessed through self-reported frequency of experiencing these feelings using 4- or 5-point Likert style responses. These questions were reproduced from the English version of the Medical Outcomes Study Short Form (SF-36), a 36-item questionnaire that measures eight dimensions of perceived health status and has been repeatedly validated for its internal reliability across multiple cohorts [19, 20]. The questions used are constituents of the mental component score, with a total score range from 3-15.

### Perceived Financial Stress

Participants were asked to rate the stress they experienced in two areas: “Hard to afford basics” and “Hard to afford medical care”. Perceived stress associated with these measures of financial hardship was assessed using 4-point Likert scale for each question. Scores were summed to create a total score range of 2-8.

### Perceived Neighborhood Stress

Participants were asked to respond using a 5-point Likert scale to 18 questions about perceptions of their neighborhood environment. A higher score on a 1-5 scale for each neighborhood perception question represented a less favorable perception of that neighborhood characteristic. Participants were also surveyed about the length of time they had lived in their neighborhood. Methods of the Neighborhood Perception Assessment have been described previously [11]. Briefly, principal components factor analyses was used to define factors from the neighborhood questionnaire data. Neighborhood questions with a loading score of 0.40 or higher were used to define the theme of each factor. Three factors were identified that were associated with neighborhood stress: (1) neighborhood violence, (2) physical environment, and (3) social cohesion. Numeric values assigned to Likert scale answers for a factor’s questions were summed to calculate a factor-related perception score. The total neighborhood perception score was the sum of factor-related perception scores and ranged from 10 to 49 with higher scores representing a more unfavorable perception of neighborhood environment.

## Data Collection and Definitions

Detailed data collection methods from study entry and follow-up have been previously reported[16]. Race and ethnicity were self-reported as non-Hispanic black, non-Hispanic white, Hispanic or other, in accordance with the categories used in the Third National Health and Nutrition Examination Survey[21]. Gender was self-reported as male or female. Other demographic information including age, insurance status, household income, achieved education and income level, marital status, family information, health behaviors and church attendance, were obtained from participants through self-report during a structured interview performed by study staff. Education was divided into 4 groups: (1) less than high school, (2), high school graduate and (3) partial college completion and (4) college graduate. Household income level was categorized into 4 groups based on annual amount (<$16,000, 16-29,999, 30-49,999, and ≥ 50,000). Health insurance status was categorized as having or not having insurance.

Body Mass Index (BMI) was calculated based on measured height and weight. Blood pressure was measured using previously described methods. Blood pressure was measured using an automatic oscillometric device (Series #52,000, Welch Allyn, Inc., Arden, North Carolina) as described previously. Hypertension was defined as systolic blood pressure ≥ 140, diastolic blood pressure ≥ 90 or the use of anti-hypertensive medications. Hypercholesterolemia was defined as use of lipid-lowering medication, fasting or non-fasting low-density lipoprotein (LDL) ≥ 160mg/dL, or total cholesterol ≥240mg/dL. Diabetes mellitus was defined as HbA1c ≥ 6.5% or prior diagnosis of diabetes or use of glucose lowering drugs. Insulin sensitivity was estimated using the Homeostasis Model Assessment of Insulin Resistance Index (HOMA-IR) calculated by multiplying fasting plasma insulin (mU/L) by fasting plasma glucose (mmol/L) and then dividing by a constant of 22.5. Cardiac biomarkers, including high-sensitivity cardiac troponin T, N-terminal pro-B-type natriuretic peptide (NT-proBNP) and high sensitivity C-reactive protein (hs-CRP) were measured using the Elecsys 2010 platform (Roche Diagnostics, Indianapolis, IN), as previously reported[22].

Study participants wore an accelerometer (Actical, Phillips Respironics, Bend, OR) for 7 consecutive days on their non-dominant wrist to measure sedentary time and physical activity[23]. Sedentary time was identified using a threshold of <100 counts per minute from 8am to 8pm to reduce the misclassification of sleep as sedentary time. Total minutes spent <100 CPM was divided by the number of days to assess average daily sedentary time. Vigorous physical activity was defined as >4000 counts/min and corresponds to ≥6 METS. Minutes spent above the threshold of 4000 CPM and between 1500 to 4000 CPM were averaged across all valid wear days to estimate the total daily duration of moderate to vigorous physical activity (MVPA).

## Clinical Outcomes

Participants were followed via an annual health survey regarding interval cardiovascular events and through quarterly tracking for hospital admissions using the Dallas-Fort Worth Hospital Council Data Initiative as described previously [16]. The study outcomes included ASCVD, Global CVD and all-cause mortality. ASCVD includes CVD death, fatal and or non-fatal MI, fatal or non-fatal stroke or coronary or peripheral revascularization. Global CVD compromises ASCVD plus hospitalization for heart failure and arial fibrillation. Events were adjudicated by a panel of cardiovascular specialists.

## Statistical Analyses

The cumulative stress score (CSS) subcomponents – PSS-4, psychosocial, financial and neighborhood stress scores -- were standardized to have equal weighting for statistical analyses and such that the average following standardization had a mean of 0 and standard deviation of 1. The cumulative stress score ranged from -5.09 to 12.61 after summation of the standardized components and was analyzed as a continuous variable. Models are reported per standard deviation unit increase.

Linear and logistic regression models were used to determine associations of the CSS with demographics, psychosocial variables, cardiac risk factors and subclinical CVD phenotypes after adjusting for age, gender, and race/ethnicity (model 1) and additionally for education and income (model 2). Cox proportional hazards models were used to determine the association of the cumulative stress score with clinical outcomes. Model 1 adjusted for age and gender. Model 2 added race/ethnicity, income, and education to Model 1. Model 3 added traditional ASCVD risk factors (hypertension, diabetes, smoking, hyperlipidemia) to Model 2. To assess for potential effect modification, testing for interaction of the cumulative stress score with age, gender, race/ethnicity, income, and education was performed, both for the linear regression and Cox proportional hazards analyses. Statistical analyses were completed using SAS (version 9.4). All P values are 2-sided with P<0.05 considered significant.

## Results

The characteristics of the study population are presented in Table 1. The overall study population consisted of 59% female, 34% White, 49% Black and 15% Hispanic/Latinx participants, with a median age of 48 years. Most of the population (52%) completed partial college education with the second highest proportion completing high school (20%).

**Table 1.** Baseline Characteristics

| Characteristics | n (%) |
| --- | --- |
| Age (median, 25/75<sup>th</sup> percentile) | 48 (40, 61) |
| Male | 1210 (41%) |
| Female | 1765 (59%) |
| Black | 1457 (49%) |
| White | 1010 (34 %) |
| Hispanic | 438 (15%) |
| Married | 1498 (50%) |
| **Education** |  |
| Less than high school | 422 (14%) |
| High school | 590 (20%) |
| Partial college | 1551 (52%) |
| Completed college | 271 (9%) |
| **Income** |  |
| Less than \$16,000/year | 477 (16%) |
| 16,000-30,000/year | 464 (16%) |
| 30,000-50,000/year | 754 (25%) |
| > 50,000/year | 1017 (34%) |
| Unemployed | 245 (8%) |
| Full time Job | 1659 (5%) |
| Health insurance | 2194 (74%) |
| **Risk Factors** |  |
| Current smoker | 688 (23%) |
| Diabetes | 446 (15%) |
| Hypercholesterolemia | 780 (26%) |
| Hypertension | 151 (51%) |
| Body mass index (median, 25/75<sup>th</sup> percentile) | 30.8 (25.7, 36.21) |
| Current drinker | 868 (30%) |

Correlation between individual stress score components and the CSS are reported in Supplementary Table 1. Psychosocial stress and PSS-4 were the most strongly correlated of the individual components (Spearman rho 0.54). Correlations of PSS-4, psychosocial, neighborhood and financial stress with the CSS were moderate to strong (rho 0.45-0.76; supplementary table 1).

CSS was higher among women, those of younger age and those with lower income and educational attainment (p <.0001 for each, Figure 1). CSS was higher among individuals self-identifying as Black or Hispanic vs White (Both P<.0001, Figure 1); however, there was no difference between Black and Hispanic participants (Figure 1). In unadjusted analyses higher CSS was strongly associated with self-report of racial discrimination, lack of health insurance, and reported last medical contact greater than one year previously (p<.0001 for each).

**Figure 1.**
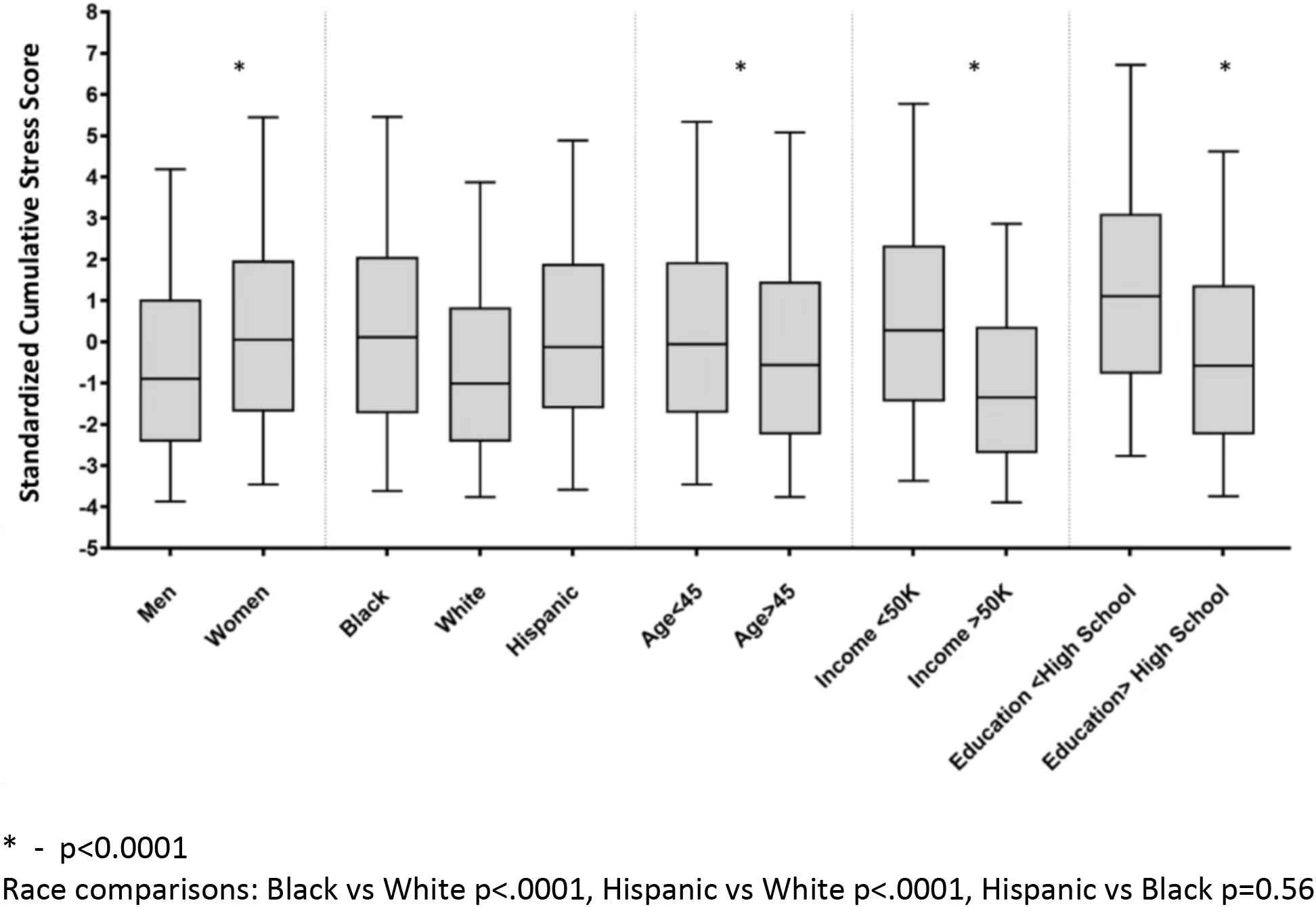
Distributions of Cumulative Stress Score in DHS-2.

**Figure 2.**
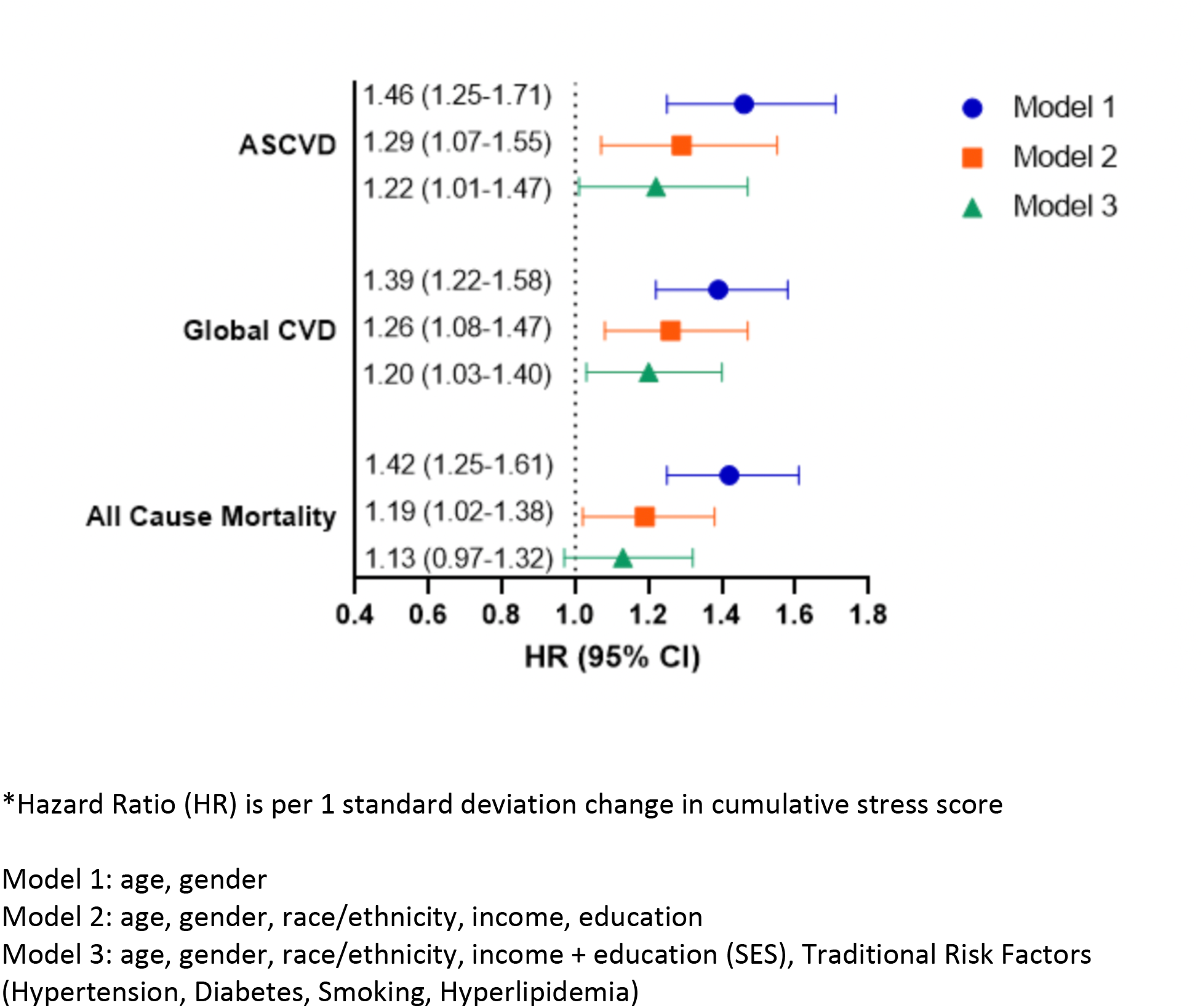
Associations of Cumulative Stress Score with Cardiovascular Outcomes and All-Cause Mortality.

### Associations of Cumulative Stress Score with CVD Risk Factors, Health Behaviors and Intermediate Phenotypes

In linear regression models adjusting for age, gender, and race/ethnicity, higher CSS was associated with higher SBP, DBP, BMI, waist circumference, HbA1c, hs-CRP, and HOMA-IR (Table 2, Model 1). There was also a significant association between CSS and accelerometer measured sedentary time and an inverse association with MVPA. All of these associations persisted after additional adjustment for income and education. The prevalence of traditional risk factors, including diabetes mellitus, current smoking, and hypertension, was higher among those with higher cumulative stress after adjustment for age, sex, and race/ethnicity though only current smoking status and hypertension achieved statistical significance after additional adjustment for education and income (Table 3). No significant association was observed between CSS and intermediate CV phenotypes such as left ventricular mass, coronary artery calcium (CAC) score or cardiovascular biomarkers such as hs-TnT or NT-proBNP (Tables 2 and 3). Associations of individual components of the CSS with health behaviors and risk factors is shown in Supplementary Table 2. Testing for interaction of the CSS by race/ethnicity, gender and age demonstrated few interactions (Supplementary table 3). There were interactions between CSS and gender for BMI and waist circumference that were further investigated with subgroup analyses and showed a stronger association of cumulative stress with obesity measures among women than men.

**Table 2.** Multivariable Associations of Cumulative Stress Score with Measured Variables.

| Variable | Model 1 |  | Model 2 |  |
| --- | --- | --- | --- | --- |
| | β-coefficient<br>(Standard Error) | P value | β-coefficient<br>(Standard Error) | P value |
| Systolic blood pressure | 1.44 (0.36) | <0.0001 | 1.09 (0.41) | <0.01 |
| Diastolic blood pressure | 0.77 (0.18) | <0.0001 | 0.73 (0.20) | <0.001 |
| Body Mass Index | 0.63 (0.14) | <0.0001 | 0.61 (0.16) | <0.001 |
| Waist Circumference | 1.59 (0.33) | <0.0001 | 1.39 (0.37) | <0.001 |
| HDL cholesterol | -0.79 (0.32) | 0.01 | -0.44 (0.36) | 0.22 |
| LDL cholesterol | -1.18 (0.71) | 0.10 | -0.11 (0.80) | 0.89 |
| Time in moderate to vigorous physical activity | -1.37 (0.07) | 0.07 | -2.12 (0.87) | 0.02 |
| Sedentary time | 8.95 (2.19) | <0.0001 | 7.93 (2.46) | <0.01 |
| Hemoglobin A1c | 0.06 (0.02) | < .01 | 0.05 (0.02) | <0.01 |
| NT-proBNP | 0.05 (0.02) | 0.01 | 0.02 (.02) | 0.30 |
| HOMA-IR | 0.33 (0.10) | <0.001 | 0.26 (0.11) | 0.04 |
| hs-CRP | 0.08 (0.03) | <0.01 | 0.07 (0.03) | 0.03 |
| hsTnT | 0.03 (0.02) | 0.08 | 0.03 (0.02) | 0.12 |
| Left ventricular mass | 0.89 (0.79) | 0.29 | 0.84 (0.79) | 0.29 |

\* $\beta$ -coefficient reflects unit of change in the variable of interest per standardized deviation change in the cumulative stress score
Model 1: age, sex, race/ethnicity
Model 2: age, sex, race/ethnicity, income, education

**Table 3.** Multivariable Associations of Cumulative Stress Score with Risk Factors and Coronary Calcium.

|  | Model 1 |  | Model 2 |  |
| --- | --- | --- | --- | --- |
| **Risk Factors and coronary calcium** | Odds Ratio (95% CI) | P value | Odds Ratio (95% CI) | P value |
| Hypertension | 1.24 (1.13-1.35) | <0.0001 | 1.18(1.07-1.30) | <0.001 |
| Current drinker | 0.93 (0.85-1.01) | 0.11 | 1.03 (0.93-1.13) | 0.62 |
| Current smoker | 1.64 (1.49-1.80) | <0.0001 | 1.46(1.31-1.62) | <0.0001 |
| Diabetes | 1.17 (1.05-1.31) | <0.01 | 1.11(0.99-1.26) | 0.09 |
| Coronary Artery Calcium ≥ 10 Agatston U | 1.11 (0.99-1.24) | 0.08 | 1.03 (0.91-1.17) | 0.64 |

Odds Ratio (OR) is per 1 standard deviation change in cumulative stress score
Model 1: age, sex, race/ethnicity
Model 2: age, sex, race/ethnicity, income, education
Coronary Artery Calcium ≥ 10 represents Coronary Artery Calcium Score of greater than or equal to 10

### Associations of Cumulative Stress Score with CVD Outcomes

Over a median of 12.4 years follow up, ASCVD occurred in 136, Global CV in 202 and All-Cause Mortality in 211 participants. In Cox proportional hazards models adjusting for age and gender, CSS was associated with higher incidence of ASCVD (HR 1.46 per standard deviation [SD] increment, 95% CI 1.25-1.71), global CVD (HR 1.39, 95% CI 1.22-1.58), and all-cause mortality (HR 1.42, 95% CI 1.25-1.61). After additional adjustment for race/ethnicity, income and education, associations were modestly attenuated but remained significant (Figure 1). In a fully adjusted model that added adjustment for traditional risk factors (Hypertension, Diabetes, Smoking, Hyperlipidemia), associations with ASCVD (HR 1.22, 95% CI 1.01-1.47) and Global CVD (HR 1.20, 95% CI 1.03-1.40) remained significant though associations with all-cause mortality were attenuated (HR 1.13, 95% CI 0.97-1.32(Figure 1). No interactions of the CSS with age, sex, race/ethnicity, income, or education were observed for any of the endpoints.

The individual stress components, PSS-4, psychosocial, financial and neighborhood stress, were each independently associated with ASCVD, Global CVD, and All-Cause Mortality in analyses adjusted for age and gender (supplementary figure 2, model 1). In the fully adjusted model, which added race/ethnicity, income, education, and traditional risk factors, neighborhood stress was independently associated with ASCVD and global CVD while psychosocial stress was associated with global CVD and all-cause mortality (supplementary figure 2, model 2).

## Discussion

In this longitudinal study of a diverse urban population-based cohort, we found that a cumulative stress score that integrated different domains of perceived stress was associated with multiple adverse cardiovascular health behaviors and CVD risk factors, as well as a significantly increased hazard of incident ASCVD and global CVD. Notably, associations with CVD outcomes persisted even after adjusting for demographics and traditional CVD risk factors. Though associations of perceived cumulative stress with risk factors and outcomes were generally similar across subgroups, the increased burden of cumulative perceived stress in women, Black and Hispanic individuals, and those with lower income and education suggests a higher attributable risk in these traditionally marginalized subgroups.

### Prior Literature

Existing literature has not directly explored associations of cumulative stress across multiple domains with incident CVD and mortality. Prior studies have demonstrated associations between increased occupational, marital, and psychosocial stress with cardiovascular disease [1–6, 24]. These studies have important limitations, however, including brief or single item assessments of stress and lack of generalizability to contemporary and diverse populations[25–27]. Moreover, studies of perceived stress and CVD outcomes have largely considered only single domain assessments. In the Jackson Heart Study population of African Americans, participants with moderate to high financial stress were found to have increased risk of incident coronary heart disease after controlling for traditional risk factors, sociodemographic factors, and access to care [9], findings concordant with those from our study. However, financial stress was not considered in the context of other stressors in that study.

Redmond et al investigated associations of perceived stress with CHD and all-cause-mortality in the REGARDS cohort[13] using the same PSS-4 included in our cumulative score. They found increased hazard for incident CHD among participants reporting the highest level of stress, with effect modification by income, such that stronger associations were seen in the low-income subgroup. While we did not find effect modification by income, we do see similar associations of higher cumulative stress score with lower income and education as was seen in REGARDS. A study by Albert et al investigated the relationship between a cumulative stress model and ideal cardiovascular health metrics as defined by the American heart Association’s 2020 strategic impact goals in the Women’s Health Study [28]. Similar to our findings, the investigators found that cumulative stress levels were higher in minoritized participants, with Hispanic, Black, and Asian participants having higher cumulative stress than White participants. Though the study did not report associations between their cumulative stress model and cardiovascular outcomes, it considered similar components of stress (financial, neighborhood, life trauma) as were included in our cumulative score. Thus, our study builds on these prior findings by demonstrating for the first-time associations of a cumulative stress assessment with important clinical CVD outcomes.

In our analyses, individual subcomponents of stress including the PSS-4 appeared to be less strongly correlated with CV outcomes than the cumulative stress score. Additionally, although in univariable analyses the individual subcomponents were all associated with incident ASCVD, Global CVD, and all-cause mortality, most of these relationships were attenuated after adjusting for sociodemographic factors and traditional risk factors. These findings suggest that assessments of individual domains of stress incompletely capture the cumulative impact of stress among diverse populations. In contrast to results from individual stress domains, CSS remained robustly associated with ASCVD and Global CVD after multivariable adjustments for demographics, income, education, and traditional risk factors. Beyond supporting the incremental value from the cumulative stress model over individual components, this finding also suggests that effects of cumulative stress on CVD may not be entirely mediated through traditional risk factors.

### Mechanisms Linking Cumulative Stress with CVD Outcomes

It is possible that our study underestimates associations of perceived stress with risk factors, as well as the contribution of risk factor burden to increased CVD risk among individuals with increased stress, due to the single time point assessments of perceived stress, health behaviors and risk factors. However, it is also plausible that increased perceived stress enhances CVD risk through alternative pathways. Emerging data on the biology of stress and adversity has expanded understanding of the pathophysiology of chronic stress and disease risk. For example, a correlation between resting amygdala activity and aortic vascular inflammation has been demonstrated, that is linked through hematopoiesis stimulated by chronic stress. This relationship has been termed the “neural hematopoietic inflammatory axis”[29] [30]. We found significant associations of the CSS with hs-CRP in our study. Other inflammatory mediators of ASCVD, such as TNF alpha, Interleukin 6 and NFKB [33] have been associated separately with psychosocial stress and depression[34, 35]. The association between these markers and cumulative stress were not directly explored in the present study but are promising candidates for future studies.

Enhanced amygdala activity and subclinical cardiovascular disease have been linked with chronic discrimination [31]. Upregulation of stress hormone secretion related to the sympathetic-adrenal-medullary or hypothalamic-pituitary-adrenal axes is another proposed mechanism for the observed association between chronic stress and CVD [32] [33]. Although we did not investigate a broad stress hormone panel, we found that HOMA-IR and HbA1c were associated with increased CSS, and it is plausible that chronic cumulative stress may impact insulin resistance and glycemic control, for example, through mechanisms such as the release of stress hormones like cortisol.

### Strengths and Limitations

Our study has multiple strengths including enrollment of a large and diverse multiethnic population, use of validated instruments to measure perceived stress across multiple domains, assessment of multiple self-reported sociodemographic factors, deep phenotyping of participants, and rigorous adjudication of incident events during follow up. In addition to examining a cumulative stress instrument with synergistic components, we also investigated the association of individual stress components with cardiovascular outcomes.

Several important limitations merit comment. First, stress and CVD risk factors were only examined at a single time point, which limits causal inferences and consideration of the dynamic nature of stress over time. Second, although the study adjusted for CVD risk factors and health behaviors there remains a risk of residual confounding from unmeasured factors associated with adverse outcomes. Finally, even with a multidimensional model of cumulative stress we were unable to assess important factors such as discrimination based on gender, racial/ethnicity, or sexual orientation, traumatic life events, or occupational stress given lack of adequate survey data for these areas.

### Clinical and Public Health Implications

Our findings suggest that screening for perceived stress and implementing efforts to reduce perceived stress levels and modify risk factors associated with stress could have favorable effects on CVD health. We found significant associations of cumulative stress with health behaviors such as smoking, sedentary time and moderate to vigorous physical activity, as well as CVD risk factors such as hypertension and obesity. Importantly, each of these behaviors and risk factors are modifiable. Thus, leveraging interventions aimed at health behavior and risk factor modification in individuals with high stress levels may be beneficial. Exercise, in particular, may have beneficial effects on both stress and CVD outcomes as it improves baroreflex dysregulation and blood pressure homeostasis observed with stress [37].

We found that associations of cumulative stress with CVD risk factors, health behaviors, and outcomes were largely similar across important subgroups defined by gender, race/ethnicity, age, income, and education. This suggests that pathophysiology of cumulative stress may be similar among diverse adults. On the other hand, the burden of chronic stress is born disproportionately by younger people, women, Black and Hispanic individuals, and those with lower education and income. For that reason, cumulative stress may be a particularly important risk marker or risk factor in minoritized individuals and those with lower socioeconomic status. In this regard, it may function like smoking, hypertension, obesity, and diabetes, which also demonstrate similar associations with outcomes but are more prevalent in individuals from minoritized groups and lower socioeconomic strata [38, 39]. It also stands to reason that high levels of stress may interfere with positive behavior change. Recognizing this differential stress burden is important when considering potential targeted efforts to mitigate stress-associated risk and to reduce health care disparities.

Targeted stress reduction or mitigation strategies such as transcendental meditation have been reported to decrease risk factors such as blood pressure ([40–42], smoking [43], and hypercholesterolemia [44], as well as reducing cardiovascular mortality in several small studies [45, 46] in diverse populations. Transcendental meditation practices, of which the goal is to achieve a wakeful hypometabolic state, aim to reduce the chronic emotional, physiological, and sympathetic arousal [40]. Community engaged programs are likely needed to co-design interventions with these stress reduction methods; these should focus on those most impacted by cumulative stressors in at risk populations and provide the social support and structure to sustain this change. Finally, social services interventions may help to in alleviate stress related to non-psychosocial stressors such as financial and neighborhood stress[47].

### Future Directions

Coping mechanisms and resilience likely modify the effects of chronic stress on cardiovascular disease. Future studies should evaluate the relationships between resilience, chronic stress, and CVD outcomes and test whether individuals with high cumulative stress may have cardiometabolic benefits from behavioral interventions that focus on strengthening of coping and resilience mechanisms. Screening for stress as a potential comorbidity in primary care settings should be studied, and interventions aimed at mitigating perceived stress explored. Our findings suggest that strategies that consider general mitigation strategies targeted to a full spectrum of stress, rather than individual stressors, may have greater yield.

## Conclusion

Cumulative stress is associated with cardiovascular risk factors as well as incident CVD even after accounting for increased risk factor burden seen with higher stress levels. These findings suggest that composite, multidimensional, assessments of perceived stress may help to identify individuals at risk for CVD who may be targeted for stress mitigation or enhanced prevention strategies. These approaches may be best focused on vulnerable populations, given the higher burden of stress in women, Black and Hispanic individuals, and those with lower income and education. Additional study is needed to replicate these findings and determine best approaches for stress and risk mitigation.

## Data Availability

The full data set is stored at the Dallas Heart Study (DHS) Data.

## Acknowledgments

The authors thank the other investigators, the staff, and the participants of the Dallas Heart Study Phase 2 for their valuable contributions.

## Author Contributions

Colby Ayers MS, Ijeoma Eleazu, and James A. de Lemos had full access to all the data in the study and take responsibility for the integrity of the data and the accuracy of the data analysis.

Concept and design: Eleazu, de Lemos.

Acquisition, analysis, or interpretation of data: de Lemos, Ayers, Eleazu

Drafting of the manuscript: Eleazu, Ayers, Navar, Salhadar, Albert, Carnethon, Brown, Ogbu-Nwobodo, Carter, Bess, Powell-Wiley, de Lemos

Critical revision of the manuscript for important intellectual content: Eleazu, Ayers, Navar, Salhadar, Albert, Carnethon, Brown, Ogbu-Nwobodo, Carter, Bess, Powell-Wiley, de Lemos

Statistical analysis: Ayers

Obtained funding: de Lemos

Supervision: Eleazu and de Lemos

## Data Availability

The full data set is stored at the Dallas Heart Study (DHS) Data.

## Code availability

The analytical methods for this study are available from the corresponding author upon appropriate request.

### Disclaimer

The information presented by the authors is their own and this material should not be interpreted as representing the official viewpoint of the University of Texas Southwestern Medical Center, the University of California, San Francisco Medical Center, Northwestern University, The Social Determinants of Obesity and Cardiovascular Risk Laboratory, Cardiovascular Branch, Division of Intramural Research, The National Heart, Lung, and Blood Institute, National Institutes of Health and The Intramural Research Program, National Institute on Minority Health and Health Disparities, National Institutes of Health]

## References

1. Rosengren, A., et al., Association of psychosocial risk factors with risk of acute myocardial infarction in 11119 cases and 13648 controls from 52 countries (the INTERHEART study): case-control study. Lancet, 2004. 364(9438): p. 953–62.

2. Richardson, S., et al., Meta-analysis of perceived stress and its association with incident coronary heart disease. Am J Cardiol, 2012. 110(12): p. 1711–6.

3. Rod, N.H., et al., Perceived stress as a risk factor for changes in health behaviour and cardiac risk profile: a longitudinal study. J Intern Med, 2009. 266(5): p. 467–75.

4. Eaker, E.D., et al., Marital status, marital strain, and risk of coronary heart disease or total mortality: the Framingham Offspring Study. Psychosom Med, 2007. 69(6): p. 509–13.

5. Muntaner, C., et al., Work organization and atherosclerosis: findings from the ARIC study. Atherosclerosis Risk in Communities. Am J Prev Med, 1998. 14(1): p. 9–18.

6. Steptoe, A., et al., Cardiovascular stress reactivity and job strain as determinants of ambulatory blood pressure at work. J Hypertens, 1995. 13(2): p. 201–10.

7. Boyle, S.H., et al., Depressive symptoms and mental stress-induced myocardial ischemia in patients with coronary heart disease. Psychosom Med, 2013. 75(9): p. 822–31.

8. Lichtman, J.H., et al., Depression as a risk factor for poor prognosis among patients with acute coronary syndrome: systematic review and recommendations: a scientific statement from the American Heart Association. Circulation, 2014. 129(12): p. 1350–69.

9. Moran, K.E., et al., Financial Stress and Risk of Coronary Heart Disease in the Jackson Heart Study. Am J Prev Med, 2019. 56(2): p. 224–231.

10. Diez Roux, A.V., et al., Area characteristics, individual-level socioeconomic indicators, and smoking in young adults: the coronary artery disease risk development in young adults study. Am J Epidemiol, 2003. 157(4): p. 315–26.

11. Powell-Wiley, T.M., et al., Relationship between perceptions about neighborhood environment and prevalent obesity: data from the Dallas Heart Study. Obesity (Silver Spring), 2013. 21(1): p. E14–21.

12. Westcott, S.K., T.T. Lewis, and M.A. Albert, Tackling Adversity and Cardiovascular Health: It is About Time. Circulation, 2023. 147(1): p. e1–e3.

13. Redmond, N., et al., Perceived stress is associated with incident coronary heart disease and all-cause mortality in low- but not high-income participants in the Reasons for Geographic And Racial Differences in Stroke study. J Am Heart Assoc, 2013. 2(6): p. e000447.

14. Victor, R.G., et al., The Dallas Heart Study: a population-based probability sample for the multidisciplinary study of ethnic differences in cardiovascular health. Am J Cardiol, 2004. 93(12): p. 1473–80.

15. Pandey, A., et al., Determinants of Racial/Ethnic Differences in Cardiorespiratory Fitness (from the Dallas Heart Study). Am J Cardiol, 2016. 118(4): p. 499–503.

16. de Lemos, J.A., et al., Multimodality Strategy for Cardiovascular Risk Assessment: Performance in 2 Population-Based Cohorts. Circulation, 2017. 135(22): p. 2119–2132.

17. Cohen, S., T. Kamarck, and R. Mermelstein, A global measure of perceived stress. J Health Soc Behav, 1983. 24(4): p. 385–96.

18. Sharp, L.K., et al., Assessing the Perceived Stress Scale for African American adults with asthma and low literacy. J Asthma, 2007. 44(4): p. 311–6.

19. McHorney, C.A., et al., The MOS 36-item Short-Form Health Survey (SF-36): III. Tests of data quality, scaling assumptions, and reliability across diverse patient groups. Med Care, 1994. 32(1): p. 40–66.

20. Ware, J.E., et al., SF-36 health survey : manual and interpretation guide. 1993, Boston: Health Institute, New England Medical Center. 1 volume (various pagings) : illustrations.

21. Zipf, G., et al., National health and nutrition examination survey: plan and operations, 1999-2010. Vital Health Stat 1, 2013(56): p. 1–37.

22. Lewis, A.A., et al., Racial Differences in Malignant Left Ventricular Hypertrophy and Incidence of Heart Failure: A Multicohort Study. Circulation, 2020. 141(12): p. 957–967.

23. Harrington, J.L., et al., Sedentary Behavior and Subclinical Cardiac Injury: Results From the Dallas Heart Study. Circulation, 2017. 136(15): p. 1451–1453.

24. Everson-Rose, S.A., et al., Chronic stress, depressive symptoms, anger, hostility, and risk of stroke and transient ischemic attack in the multi-ethnic study of atherosclerosis. Stroke, 2014. 45(8): p. 2318–23.

25. Iso, H., et al., Perceived mental stress and mortality from cardiovascular disease among Japanese men and women: the Japan Collaborative Cohort Study for Evaluation of Cancer Risk Sponsored by Monbusho (JACC Study). Circulation, 2002. 106(10): p. 1229–36.

26. Nabi, H., et al., Increased risk of coronary heart disease among individuals reporting adverse impact of stress on their health: the Whitehall II prospective cohort study. Eur Heart J, 2013. 34(34): p. 2697–705.

27. Ohlin, B., et al., Chronic psychosocial stress predicts long-term cardiovascular morbidity and mortality in middle-aged men. Eur Heart J, 2004. 25(10): p. 867–73.

28. Albert, M.A., et al., Cumulative psychological stress and cardiovascular disease risk in middle aged and older women: Rationale, design, and baseline characteristics. Am Heart J, 2017. 192: p. 1–12.

29. Tawakol, A., et al., Relation between resting amygdalar activity and cardiovascular events: a longitudinal and cohort study. Lancet, 2017. 389(10071): p. 834–845.

30. Stiekema, L.C.A., et al., The maturation of a ’neural-hematopoietic’ inflammatory axis in cardiovascular disease. Curr Opin Lipidol, 2017. 28(6): p. 507–512.

31. Powell-Wiley, T.M., et al., Chronic Stress-Related Neural Activity Associates With Subclinical Cardiovascular Disease in a Community-Based Cohort: Data From the Washington, D.C. Cardiovascular Health and Needs Assessment. Front Cardiovasc Med, 2021. 8: p. 599341.

32. Baumer, Y., et al., By what molecular mechanisms do social determinants impact cardiometabolic risk? Clinical Science, 2023. 137(6): p. 469-+.

33. Lagraauw, H.M., J. Kuiper, and I. Bot, Acute and chronic psychological stress as risk factors for cardiovascular disease: Insights gained from epidemiological, clinical and experimental studies. Brain Behav Immun, 2015. 50: p. 18–30.

34. Vaccarino, V., et al., Depression, inflammation, and incident cardiovascular disease in women with suspected coronary ischemia: the National Heart, Lung, and Blood Institute-sponsored WISE study. J Am Coll Cardiol, 2007. 50(21): p. 2044–50.

35. Kop, W.J., et al., Association between depressive symptoms and fibrosis markers: the Cardiovascular Health Study. Brain Behav Immun, 2010. 24(2): p. 229–35.

36. Cummings, D.M., et al., Consequences of Comorbidity of Elevated Stress and/or Depressive Symptoms and Incident Cardiovascular Outcomes in Diabetes: Results From the REasons for Geographic And Racial Differences in Stroke (REGARDS) Study. Diabetes Care, 2016. 39(1): p. 101–9.

37. Mameletzi, D., et al., Effects of long-term exercise training on cardiac baroreflex sensitivity in patients with coronary artery disease: a randomized controlled trial. Clin Rehabil, 2011. 25(3): p. 217–27.

38. Hamad, R., et al., Association of Low Socioeconomic Status With Premature Coronary Heart Disease in US Adults. JAMA Cardiol, 2020. 5(8): p. 899–908.

39. Havranek, E.P., et al., Social Determinants of Risk and Outcomes for Cardiovascular Disease: A Scientific Statement From the American Heart Association. Circulation, 2015. 132(9): p. 873–98.

40. Wenneberg, S.R., et al., A controlled study of the effects of the Transcendental Meditation program on cardiovascular reactivity and ambulatory blood pressure. Int J Neurosci, 1997. 89(1-2): p. 15–28.

41. Alexander, C.N., et al., Transcendental meditation, mindfulness, and longevity: an experimental study with the elderly. J Pers Soc Psychol, 1989. 57(6): p. 950–64.

42. Schneider, R.H., et al., A randomized controlled trial of stress reduction in African Americans treated for hypertension for over one year. Am J Hypertens, 2005. 18(1): p. 88–98.

43. Schneider, R.H., et al., Stress Reduction in the Prevention of Left Ventricular Hypertrophy: A Randomized Controlled Trial of Transcendental Meditation and Health Education in Hypertensive African Americans. Ethn Dis, 2019. 29(4): p. 577–586.

44. Castillo-Richmond, A., et al., Effects of stress reduction on carotid atherosclerosis in hypertensive African Americans. Stroke, 2000. 31(3): p. 568–73.

45. Schneider, R.H., et al., Long-term effects of stress reduction on mortality in persons > or = 55 years of age with systemic hypertension. Am J Cardiol, 2005. 95(9): p. 1060–4.

46. Schneider, R.H., et al., Stress reduction in the secondary prevention of cardiovascular disease: randomized, controlled trial of transcendental meditation and health education in Blacks. Circ Cardiovasc Qual Outcomes, 2012. 5(6): p. 750–8.

47. Frieden, T.R., A framework for public health action: the health impact pyramid. Am J Public Health, 2010. 100(4): p. 590–5.

